# First-in-human imaging with [^89^Zr]Zr-DFO-SC16.56 anti-DLL3 antibody in patients with high-grade neuroendocrine tumors of the lung and prostate

**DOI:** 10.1101/2024.01.10.24301109

**Authors:** Salomon Tendler, Mark P. Dunphy, Matthew Agee, Joseph O’Donoghue, Rania G. Aly, Noura J. Choudhury, Adam Kesner, Assen Kirov, Audrey Mauguen, Marina K. Baine, Heiko Schoder, Wolfgang A Weber, Natasha Rekhtman, Serge K. Lyashchenko, Lisa Bodei, Michael J. Morris, Jason S. Lewis, Charles M. Rudin, John T. Poirier

**Author notes:** Equal contribution (Salomon Tendler, Mark P. Dunphy).

## Abstract

**Background:** Delta-like ligand 3 (DLL3) is aberrantly expressed on the cell surface in many neuroendocrine cancers including small cell lung cancer (SCLC) and neuroendocrine prostate cancer (NEPC). Several therapeutic agents targeting DLL3 are in active clinical development. Molecular imaging of DLL3 would enable non-invasive diagnostic assessment to inform the use of DLL3-targeting therapeutics or to assess disease treatment response.

**Methods:** We conducted a first-in-human immuno-positron emission tomography (immunoPET) imaging study of [^89^Zr]Zr-DFO-SC16.56, composed of the anti-DLL3 antibody SC16.56 conjugated to desferrioxamine (DFO) and the positron-emitting radionuclide zirconium-89, in 18 patients with neuroendocrine cancers. An initial cohort of three patients received 1–2 mCi of [^89^Zr]Zr-DFO-SC16.56 at a total mass dose of 2·5 mg and underwent serial PET and computed tomography (CT) imaging over the course of one week. Radiotracer clearance, tumor uptake, and radiation dosimetry were estimated. An expansion cohort of 15 additional patients were imaged using the initial activity and mass dose. Retrospectively collected tumor biopsies were assessed for DLL3 by immunohistochemistry (IHC) (n = 16).

**Findings:** Imaging of the initial 3 SCLC patients demonstrated strong tumor-specific uptake of [^89^Zr]Zr-DFO-SC16.56, with similar tumor: background ratios at days 3, 4, and 7 post-injection. Serum clearance was bi-phasic with an estimated terminal clearance half-time of 119 h. The sites of highest background tracer uptake were blood pool and liver. The normal tissue receiving the highest radiation dose was liver; 1·8 mGy/MBq, and the effective dose was 0.49 mSv/MBq. Tumoral uptake varied both between and within patients, and across anatomic sites, with a wide range in SUVmax (from 3·3 to 66·7). Tumor uptake by [^89^Zr]Zr-DFO-SC16.56 was associated with protein expression in all cases. Two non-avid DLL3 NEPC cases by PET scanning demonstrated the lowest DLL3 expression by tumor immunohistochemistry. Only one patient had a grade 1 allergic reaction, while no grade ≥2 adverse events noted.

**Interpretation:** DLL3 PET imaging of patients with neuroendocrine cancers is safe and feasible. These results demonstrate the potential utility of [^89^Zr]Zr-DFO-SC16.56 for non-invasive in vivo detection of DLL3-expressing malignancies.

**Funding:** Supported by NIH R01CA213448 (JTP), R35 CA263816 (CMR), U24 CA213274 (CMR), R35 CA232130 (JSL), and a Prostate Cancer Foundation TACTICAL Award (JSL), Scannell foundation. The Radiochemistry and Molecular Imaging Probes Core Facility is supported by NIH P30 CA08748.

## Background

Delta-like ligand 3 (DLL3) is a cell surface protein that was nominated as a selective marker of high-grade neuroendocrine tumors, notable for the absence of cell surface expression in normal adult tissues.(1) The tumor selectivity of DLL3 expression has prompted the development of several DLL3-targeting agents, which are currently under clinical investigation in patients with a variety of high-grade neuroendocrine cancers.(2, 3) DLL3 expression is observed in >75% of small cell lung cancer (SCLC); expression is less common among lung adenocarcinoma with neuroendocrine features (54%) and atypical carcinoid tumors (24%).(4) Neuroendocrine prostate cancer (NEPC) is a histologic subtype of prostate cancer that most often arises from castration-resistant prostate adenocarcinoma (CRPC) with acquired resistance to androgen suppression.(5) DLL3 is expressed in >75% of NEPC, in contrast to infrequent expression in mCRPC lacking neuroendocrine features (12·5%), localized PC (0·5%) and benign prostate (0%).(6) The pathologic, clinical, and molecular features of NEPC, which include the loss of tumor suppressor genes *TP53* and *RB1* and the expression of neuroendocrine markers, closely resemble those of *de novo* SCLC.(7) Recent studies have emphasized the increasing incidence of NEPC: 15–20% of advanced CRPC cases transform into NEPC.(8) The rising incidence of NEPC might be explained in part by development and use of increasingly effective androgen receptor (AR) pathway inhibitors, reducing other mechanisms of tumor escape.(8) Since mCRPC is a heterogeneous disease, biopsies of individual lesions poorly represent the full spectrum of potential neuroendocrine features within a given patient, and therefore the incidence of NEPC is likely to be underestimated.

While transformed NEPC may be suspected in patients who develop rapidly progressive disease, visceral metastases, low ratios of PSA values to tumor burden, unusual metastatic sites, or stereotyped patterns of metastasis, there remains a lack of consensus on the optimal timing or nature of diagnostic interventions to evaluate potential NEPC transformation.(9) Accurate diagnosis is further complicated by tumor heterogeneity: CRPC exists on a spectrum in the evolution to NEPC and can encompass both pure and mixed-histology tumors.(10) DLL3 expression has previously been assessed by immunohistochemistry (IHC) performed on biopsy material obtained from primary tumor or a metastatic lesion at the time of diagnosis.

SC16.56 is a humanized DLL3-specific IgG1 monoclonal antibody originally developed as part of an antibody-drug conjugate tested in clinical trials, which we have repurposed as a non-invasive immunoPET imaging probe.(11–16) In this report, we describe the results of a pilot study using [^89^Zr]Zr-DFO-SC16.56 to image disease in patients with histologically-verified neuroendocrine tumors, including patients with SCLC, NEPC, lung adenocarcinoma with neuroendocrine features, and atypical carcinoid. We investigated the tolerability and feasibility of immunoPET/CT imaging of DLL3 expression using [^89^Zr]Zr-DFO-SC16.56, with parallel assessment of tumor DLL3 expression by IHC, as well as assessment of the pharmacokinetics, biodistribution, metabolism, and radiation dosimetry of this radioimmunoconjugate.

## Materials and Methods

### Study Population

The study was an open-label, nonrandomized, first-in-human study conducted at Memorial Sloan Kettering Cancer Center (MSKCC), with the approval of the institutional review board (NCT04199741). The study was conducted under the auspices of an Investigational New Drug application held by the institution and approved by the US Food and Drug Administration. All participants provided written informed consent. The study was conducted in accordance with the Declaration of Helsinki.

Subjects were enrolled from January 2020 until January 2023 and patient inclusion criteria included a minimum age of 18, with histologically verified, neuroendocrine-derived malignancies, with at least one tumor lesion measuring ≥ 1.5 cm on a conventional radiological scan such as body CT, 2-deoxy-2-[fluorine-18]fluoro-D-glucose (^18^F-FDG) PET scan, and/or magnetic resonance imaging (MRI) of the brain. The scans had to be obtained within 8 weeks prior to enrollment. Other inclusion criteria included ECOG performance status 0–2, and serum renal and hepatic function test values <1·5–5-fold greater than the lab-specific upper limit of normal. Exclusion criteria included pregnancy, breastfeeding, and acute psychiatric illnesses. (**Supplementary Table 1**).

### Study design

The study included two cohorts, with Cohort 1 including three SCLC patients who underwent serial PET/CT imaging, whole-body probe measurements, and blood sampling according to previously described methods.(17, 18) These patients received a single injection of e (nominally 2 mCi of ^89^Zr) and multiple scans were acquired between 1 hour and 7 days post-injection as indicated in **Figure 1**, with no co-injection of unconjugated SC16.56. The trial design included an optional cohort to also receive co-injected unconjugated SC16.56, but this was not pursued as the Cohort 1 demonstrated satisfactory imaging characteristics. The aim of the initial phase of the study was to evaluate the pharmacokinetics, biodistribution, and safety of [^89^Zr]Zr-DFO-SC16.56 and to estimate radiation doses to normal tissues.

**Figure 1:**
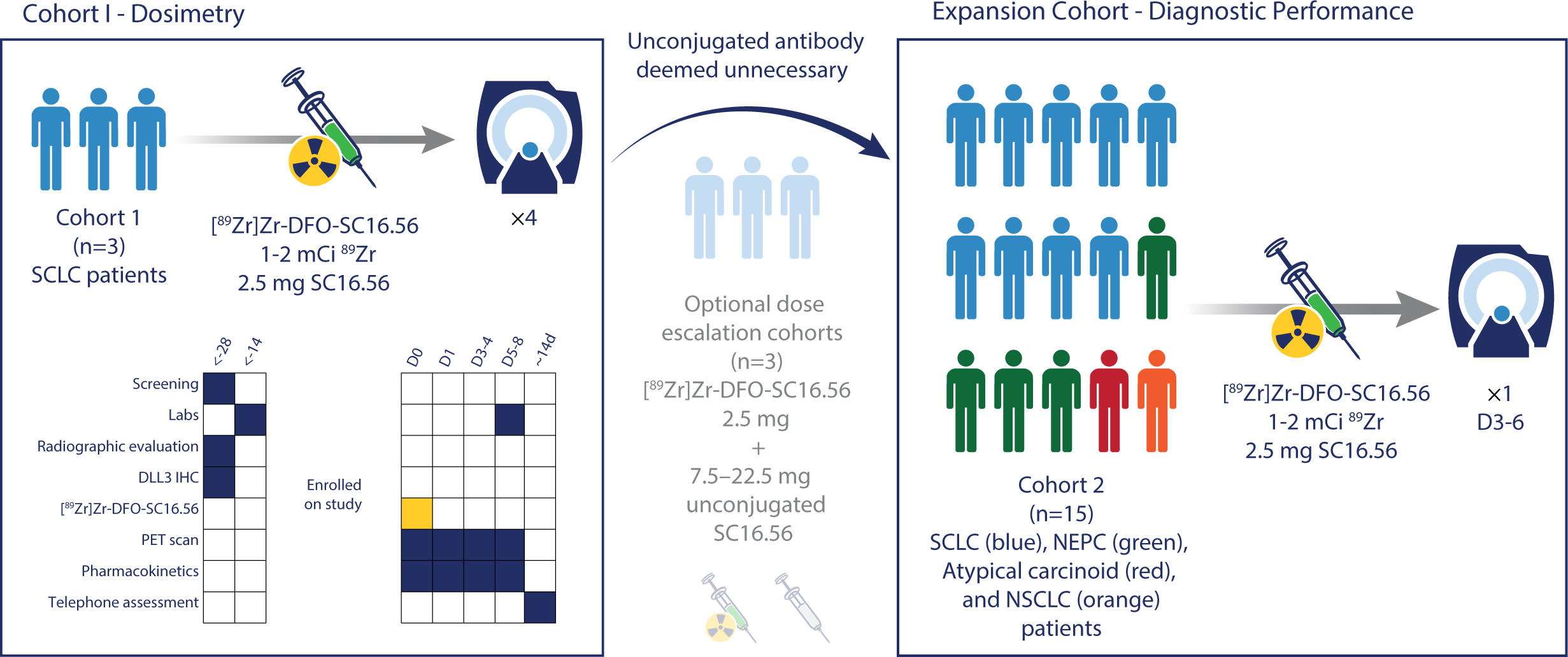
The design of the first-in-human 89Zr-DFO-SC16.56 open-label, nonrandomized study. Cohort I included three patients with SCLC, with four imaging time points selected: 1 hour – D7 pi (left). The pre-planned dose-escalation of unconjugated SC16.56 was deemed unnecessary (middle). The expansion cohort included 15 patients with different histologies that were injected with a single dose of 89Zr-DFO-SC16.56 and imaged between D3-D6 (right).

The expansion cohort included 15 patients that received a single injection of the radioimmunoconjugate followed by a single PET/CT scan three to six days later, based on observed optimal scan interval in Cohort 1. The aims of the expansion cohort included obtaining further safety and performance data on the diagnostic tracer in a larger number of patients with different cancer diagnoses and exploring the potential association between tumor uptake of the tracer and intratumoral DLL3 protein expression.

### Radiopharmaceutical preparation

SC16.56 antibody was generously provided by AbbVie Inc (Chicago, IL). The radioimmunoconjugate [^89^Zr]Zr-DFO-SC16.56was manufactured at the MSK Radiochemistry and Molecular Imaging Probes Core Facility in compliance with the requirements specified in the Chemistry, Manufacturing, and Controls section of FDA IND #145,626. SC16.56 was conjugated with a bifunctional chelator, p-SCN-deferoxamine (Macrocyclics, Plano, TX), and subsequent radiolabeling with ^89^Zr-oxalate (3D Imaging LLC, Little Rock, AR). The conjugation to DFO involved buffer exchanging the SC16.56 antibody from its native formulation into 0.1 molar (M) sodium bicarbonate solution using tangential flow filtration with a 30kDa cutoff membrane size. Once the initial buffer exchange had been completed, 6M of DFO per mole of antibody was added and allowed to react at 37°C for 90 minutes. Following incubation, tangential flow filtration was applied again to remove any unreacted DFO and to buffer exchange the DFO-SC16.56 into 1M ammonium acetate (pH 7 solution). The prepared DFO-SC16.56 precursor material was sterile filtered and vialed in 3 mg aliquots into sterile polypropylene tubes and stored at <60°C until use. The conjugation protein yield was ∼60% and the obtained number of chelates per antibody was < 3.

Radiolabeling of DFO-SC16.56 with ^89^Zr was performed by buffering the ^89^Zr in 1M oxalic acid precursor solution to pH 7 with 2M sodium carbonate solution and 1M ammonium acetate solution, followed by the addition of ∼2·5 mg of DFO-SC16.56 and incubation at ambient temperature for 50 minutes. After incubation, the [^89^Zr]Zr-DFO-SC16.56 was purified using size-exclusion desalting gel PD-10 columns (GE Lifesciences Inc, Westborough, MA) into 5mL of 0·5% gentisic acid in 0·25M sodium acetate solution.

All batches of [^89^Zr]Zr-DFO-SC16.56 underwent quality control testing prior to patient administration. The mean radionuclide incorporation in the [^89^Zr]Zr-DFO-SC16.56 final drug product was > 90%, as measured by radio-TLC, and the antibody monomer content in the final [^89^Zr]-DFO-SC16.56 drug product was consistently >90%, as measured by size exclusion-high-performance liquid chromatography (**Supplementary Figure 1**).

Patients received the radioimmunoconjugate by intravenous (IV) infusion over 10 minutes. No premedication was administered. Patients were advised to monitor for any potential symptoms up to 30 days post-injection and the subjects were evaluated at each imaging time point. All observed toxicities were graded per the common terminology criteria for adverse events version 5.0 (CTCAEv5).(19)

### Whole body counting, blood sampling and immunoPET/CT imaging

For Cohort 1, whole-body (WB) clearance was estimated by serial measurements of count-rate using a 12·7-cm-thick NaI (Tl) scintillation detector at a fixed distance (3 m) from the patient. Duplicate anterior and posterior measurements were made and background-corrected geometric mean values were used as the representative metric. WB counts were acquired immediately before and after first void and at the times of PET/CT scanning. Additionally, serial blood samples were drawn at 15, 30, 60, and 120 min and at the times of imaging. Serum samples were counted in a gamma-well-type detector (Wallac Wizard 1480 γ-counter; Perkin Elmer) to determine the percentage of injected activity per liter of serum. Mono- and bi-exponential functions were fitted to the whole-body and serum clearance data, respectively.

[^89^Zr]Zr-DFO-SC16.56 PET/CT images from skull vertex to proximal thigh were acquired on a single Discovery 710™ PET-CT imaging system (General Electric, USA). CT images were acquired with 140 kVp, with current scaled according to body weight up to 80 mA, a pitch of 1·75:1, reconstructed slice thickness of 3·75 mm, and a 0.8 s per rotation. PET data was acquired typically for 45–60 minutes in approximately 6–7 bed positions and reconstructed by ordered subset expectation maximization (OSEM; 2-iterations, 16-subsets) into a 128 × 128 matrix with all manufacturer corrections applied viz CT-based attenuation, scatter, time-of-flight (TOF), and point-spread-function (PSF).

In addition, PET images were reconstructed using GE’s proprietary Q.clear technology into a 256 x 256 matrix. Q.Clear uses a block sequential regularized expectation maximization (BSREM) algorithm that allows each voxel to achieve full convergence, potentially providing more accurate activity quantification. The β factor that controls the relative strength of the penalty function, and is the algorithm’s only adjustable parameter, was set within the range 1600–3000 to optimize image quality, based on prior work.(20) [^89^Zr]Zr-DFO-SC16.56 PET/CT images were analyzed by an experienced nuclear medicine physician using commercial display software (HERMES Gold4.4-B, HERMES Medical Solutions AB, Stockholm, Sweden; and AW Centricity Imaging-PACS/AW Suite, GE Healthcare Integrated IT Solutions, Barrington, IL). The DLL3 immunoPET/CT images were analyzed together with the most recent radiological scans: [^18^F]FDG PET/CT or CT of thorax/abdomen, or CT/MRI brain scans, as appropriate.

For dosimetry purposes, whole-organ time-integrated activity coefficients (TIAC) were derived from image-based AUC estimates and used as input data for OLINDA/EXM v.2·1, an FDA-approved radiation dosimetry software package.(21)

### DLL3 expression as determined by Immunohistochemistry

Tumor tissue biopsied at any time prior to investigational imaging was assessed for DLL3 expression by IHC when feasible, using a standardized Ventana platform assay (clone SP347, Ventana, Roche, Tucson, AZ, USA)(22). Semi-quantitative assessment of DLL3 expression was based on estimating the percentage of positive tumor cells (tumor proportion score, range 0–100%) multiplied by staining intensity (range 0–3) to generate an H-score (range, 0–300).(23) All staining assessments were performed by a professional pathologist.

### Tumor analytics

Tumor standard uptake values (SUV) measured from reconstructed PET images using Q.clear, and were expressed in terms of SUVmax. SUV represents the PET-measured tissue uptake as the fraction of the injected tracer dose per mL of tissue, normalized to patient mass. SUVmax was used because it reflects the highest lesion SUV and is relatively agnostic to the placement of the volume of interest.

For quantitative tumor analyses, the tumors with the highest SUV were selected from each patient. Tumors were dichotomized as avid or not avid based on appearance on the immunoPET/CT images performed in the interval of D3–D6. Tumors with tracer retention greater than blood pool were considered avid.

## Results

The study included 18 patients including patients with histology-confirmed SCLC (n = 12), NEPC (n = 4), NSCLC with neuroendocrine features (n = 1), and atypical carcinoid (n = 1). Demographics of the entire cohort are described in **Table 1**. The age range of the cohort was 23–81 years.

**Table 1:** Demographics and tumor characteristics of Cohort 1 (left) and Expansion cohort (right).

| **Table 1** | **Cohort 1**<br>n=3 | **Expansion cohort**<br>n=15 |
| --- | --- | --- |
| **Median age at time of enrollment**<br>(y, range) | **64 (23-64)** | **64 (44-81)** |
| **Sex** |  |  |
| Female | **1 (33.3)** | **5 (33.3)** |
| Male | **2 (66.7)** | **10 (66.7)** |
| **Diagnosis** |  |  |
| SCLC | **3 (100)** | **9 (60)** |
| NEPC | **0** | **4 (26.7)** |
| Atypical Carcinoid/NSCLC | **0** | **2 (13.3)** |
| **Toxicity** |  |  |
| CTCAE Grade 0 | **2 (66.7)** | **15 (100)** |
| CTCAE Grade 1 | **1 (33.3)** | **0** |
| CTCAE Grade $\geq 2$ | **0** | **0** |
| **Imaging time points DLL3 PET** |  |  |
| 1 hr | **3 (100)** | **0** |
| D1 | **3 (100)** | **0** |
| D3 | **2 (66.7)** | **6 (40)** |
| D4 | **0** | **3 (20)** |
| D5 | **0** | **5 (33.3)** |
| D6 | **0** | **1 (6.7)** |
| D7 | **3 (100)** | **0** |
| **TNM Stage prior to DLL3 PET** |  |  |
| Stage I-III | **0** | **0** |
| Stage IV | **3 (100)** | **15 (100)** |
| **Number of systemic lines of therapy prior to DLL3 PET** |  |  |
| 1-2 | **1 (33.3)** | **7 (46.7)** |
| 3-4 | **0** | **5 (33.3)** |
| 5-6 | **1 (33.3)** | **1 (6.7)** |
| > 6 | **1 (33.3)** | **2 (13.3)** |

### Safety and Tolerability

Injections of the diagnostic tracer was well tolerated, with only one grade 1 reaction (patient #3) noted across Cohort 1 during the monitoring period. This patient had a rash localized to the chest that started the day after the injection of the radioimmunoconjugate. The rash required topical diphenhydramine, before subsiding a few weeks later. There were no recorded adverse events observed in the expansion cohort. **Table 1**

### Serum kinetics and biodistribution in Cohort 1

Tumor-specific uptake of [^89^Zr]Zr-DFO-SC16.56 was observed in all three initial patients with SCLC imaged on study. Serum clearance was bi-phasic with an apparent terminal clearance half-time of 119 h (standard deviation 31) (**Figure 2A**). **Figure 2B** shows the temporal evolution of [^89^Zr]Zr-DFO-SC16.56 biodistribution over a 7-day period in the second patient with SCLC imaged. The primarily excretion pathway appeared to be hepatobiliary with little to no evident renal excretion. Tumor-specific uptake was evident between D3 and D7-post-injection (for eg. a metastasis in right hepatic lobe had a SUVmax at D3 of 15·0 and 21·8 at D7). Given the satisfactory performance of the radioimmunoconjugate in Cohort 1, the optional exploration of the use of cold blocking antibody to reduce background was not pursued, in favor of gaining further safety and feasibility data with [^89^Zr]Zr-DFO-SC16.56 alone.

**Figure 2:**
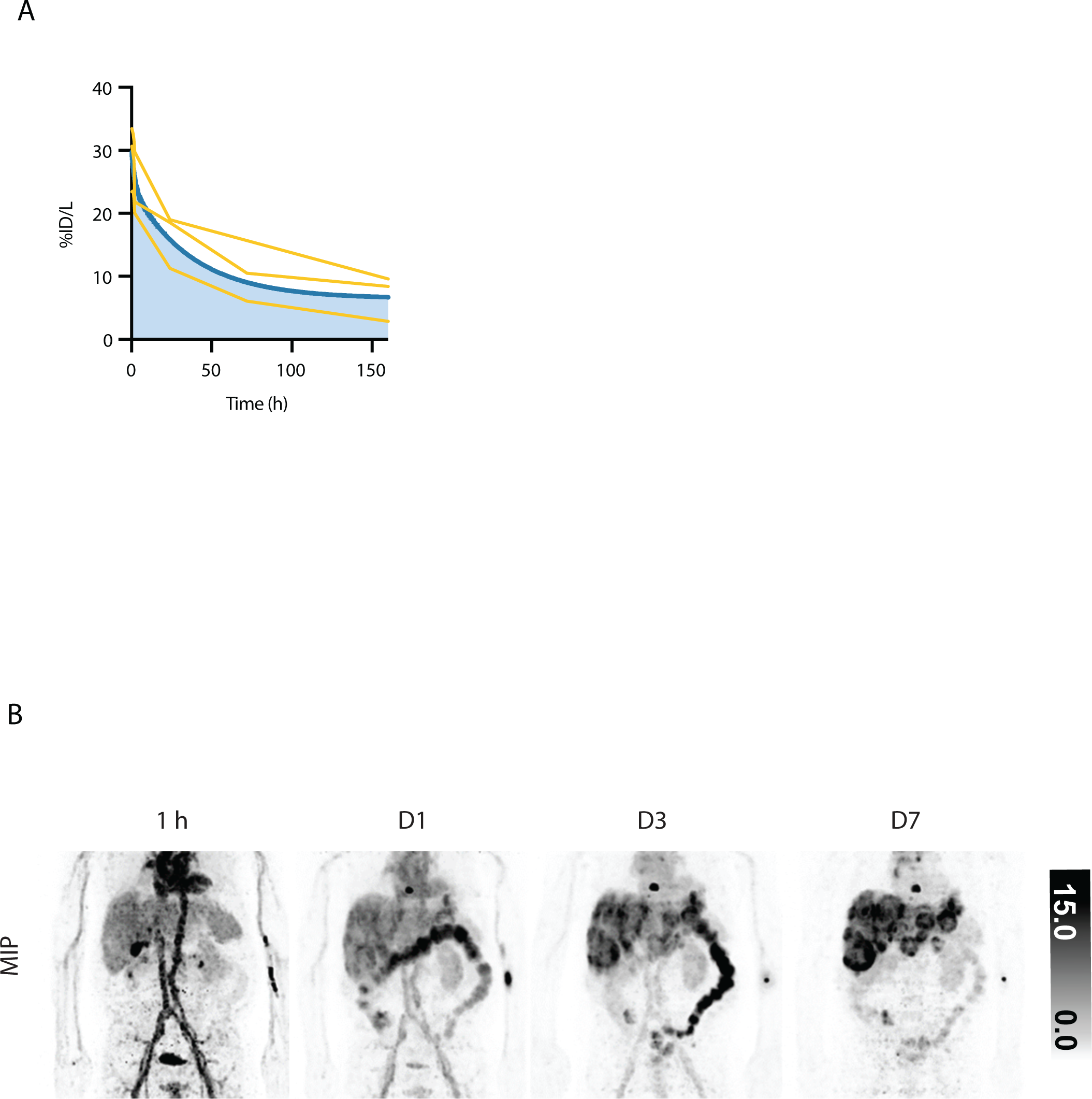
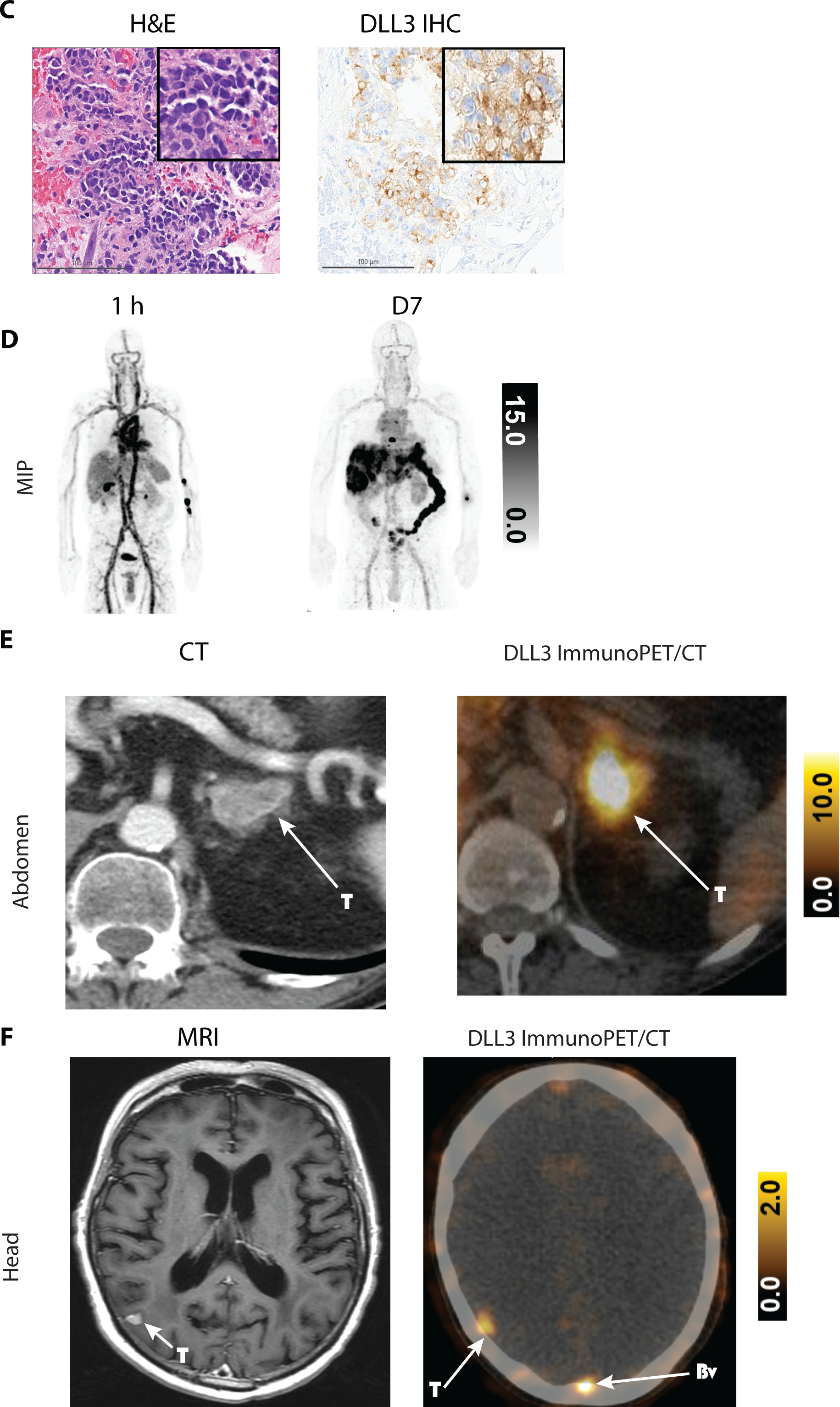
(A) Serum clearance of [89Zr]Zr-DFO-SC16.56 based on gamma well counter measurements and expressed as percentage of administered dose per liter (%ID/L). Yellow curves indicate individual patients, and the blue curve indicates the average bi-exponential clearance curve. **(B)** SCLC patient from Cohort 1, showing the temporal evolution of the biodistribution. Activity gradually clears from the circulation and tumor uptake becomes more apparent. Images also show the primarily hepatobiliary excretion pathway with only minor activity seen in the urinary bladder at the earliest time point. SUV display scale, pixels in PET imagery with SUV 0.0 appear white and pixels with SUV ≥ 15.0 appear black. The same patient (#2) had metastatic lesions in the liver, adrenal glands, and brain. The biopsied left adrenal gland metastasis **(C)** showed corresponding high DLL3 protein expression, which corresponded to uptake of ^89^Zr-DFO-SC16.56 on the DLL3 PET at D7 post-injection **(D and E)**. In addition, a single brain metastasis was also shown to be DLL3-avid on the radiological scan, which corresponded to the MRI **(F)**, with blood uptake of the tracer also observed in the posterior occipital fossa (Bv). SUV display scale, pixels in PET imagery with SUV 0.0 appear white and pixels with SUV ≥ 15.0 appear black.

Radiation doses to normal tissues from [^89^Zr]Zr-DFO-SC16.56 were estimated for the 3 patients in Cohort 1 (**Supplementary Table 2**). The normal tissue receiving the highest radiation dose was liver; 1·83 (SD 0·36) mGy/MBq, and the effective dose was 0·49 (SD 0·10) mSv/MBq. Although there were few patients for whom dosimetry was performed, the dose estimates were similar to previously reported values for [^89^Zr]Zr-pertuzumab, a diagnostic tracer for human epidermal growth factor receptor 2 (HER2)-positive tumors (**Supplementary Figure 3**).(18)

### Uptake of [^89^Zr]Zr-DFO-SC16.56 in metastatic lesions for Cohort 1

Figure 2C-F illustrates a patient (#2) from Cohort 1 diagnosed with SCLC with multiple metastatic lesions in several organs, including pericardium, liver, adrenal gland, and brain. The patient had been treated with multiple chemo- and immunotherapies prior to the imaging study with [^89^Zr]Zr-DFO-SC16.56. The serum clearance dropped from 22·3% ID/L (1-hour post-injection) to 2.8 % ID/L (D7). PET imaging showed that most metastatic lesions were DLL3-avid, with the highest uptake observed in a T8 vertebral metastasis (D3 SUVmax 54·4; D7 SUVmax 29·8). High uptake was also seen in a previously biopsied left adrenal gland metastasis (D7 SUVmax 14.6) with high DLL3 protein expression on IHC (H-score 200). The patient was subsequently enrolled onto a DLL3-directed T-cell engager therapeutic protocol.

### Correlation between tumor uptake of [^89^Zr]Zr-DFO-SC16.56 with expression of DLL3 as determined by immunohistochemistry for the entire cohort

To evaluate DLL3 protein expression by IHC staining were available either from diagnosis or subsequently in most cases (n = 16). Two cases had a IHC H-score < 50, and those two subjects also had no corresponding tumoral uptake of the [^89^Zr]Zr-DFO-SC16.56. The other 14 subjects had an H-score ≥ 150, with only one of these cases (#7) demonstrating no tumoral uptake of the diagnostic tracer.

### Expansion cohort: Tumor uptake of [^89^Zr]Zr-DFO-SC16.56 across multiple metastatic lesions and subgroups of neuroendocrine cancers in the expansion cohort

[^89^Zr]Zr-DFO-SC16.56 was further evaluated in a 15-patient expansion cohort. As summarized in **Table 2** and shown in **Supplementary Figure 2**, the tumoral take of the radioimmunoconjugate varied both between and within patients, with a wide range in SUVmax (from 3.3 to 66.7), and metastatic lesion sites (including brain, adrenal, lymph nodes, pleural, and skeletal metastasis).

**Table 2:** Patient characteristics for each individual patient included in the study.

| **Pt** | **DLL3-avid** | **SUVmax**<br>**Highest DLL3-avid lesion**<br>**&**<br>**(Anatomic location of most avid lesion)** | **Blood uptake**<br>**from D3-D6**<br>**images**<br>**(SUVaverage)** | **Tumor: Liver**<br>**ratios**<br>**(SUVmax: SUVmean)** | **Biopsy location**<br>**&**<br>**IHC H-Score** |
| --- | --- | --- | --- | --- | --- |
| 1 | Yes | 3.5<br>(Brain) | N/A | N/A | Brain met<br>(195) |
| 2 | Yes | 36.4<br>(Vertebrae) | 4.5 | 8.4 | Left adrenal<br>(200) |
| 3 | Yes | 12.8<br>(Lymph node) | N/A* | N/A | Pleura<br>(280) |
| 4 | No | 4.0 | 6.1 | N/A | Lung<br>(30) |
| 5 | Yes | 66.7<br>(Lymph node) | 10.7 | 9.3 | Lymph node<br>(300) |
| 6 | Yes | 50.5<br>(Liver) | 6.9 | 6.7 | N/A |
| 7 | No | 4.0 | 6.7 | N/A | Lymph node<br>(185) |
| 8 | Yes | 18.4<br>(Liver) | 7.6 | 3.2 | Liver<br>(290) |
| 9 | Yes | 9.5<br>(Lymph node) | 5.6 | 2.8 | Lymph Node<br>(260) |
| 10 | Yes | 16.0<br>(Lymph node) | 12.3 | 1.6 | Lung<br>(280) |
| 11 | Yes | 12.8<br>(Vertebrae) | 2.1 | 10.0 | Lymph Node<br>(150) |
| 12 | No | 3.3 | 3.3 | N/A | Liver<br>(35) |
| 13 | Yes | 27.4<br>(Lymph node) | 3.9 | 9.6 | Lymph node<br>(275) |
| 14 | Yes | 9.6<br>(Vertebrae) | 1.8 | 1.8 | Liver<br>(295) |
| 15 | Yes | 5.9<br>(Pelvis) | 4.9 | 1.2 | Lung<br>(160) |
| 16 | Yes | 25.7<br>(Lymph node) | 1.9 | 2.0 | N/A |
| 17 | Yes | 4.5<br>(Lung) | 2.1 | 2.1 | Lung<br>(190) |
| 18 | Yes | 10.6<br>(Liver) | 0.8 | 4.9 | Lymph node<br>(175) |

Abbreviations: DLL3: delta-like ligand 3, IHC: immunohistochemistry, Nr: Number, pi: post-injection, Pt: patient, N/A: not applicable, SUV: standardized uptake value, \*patient did not have D3-D6 imaging performed.

The results from this cohort highlighted that a single immunoPET/CT scan at 3-6 days post-administration could delineate DLL3-avid tumors in most cases (12/15) including metastases located in lungs, lymph nodes, liver, skeletal system, adrenal glands, and brain. The highest background uptake of the diagnostic tracer for two patients with D3 imaging from Cohort 1 and the entire expansion cohort (n=17) was found in the blood pool (SUVaverage 4·93) and liver (SUVaverage 5·39). (**Supplementary Table 3**)

Furthermore, the uptake of the tracer in the patients from the expansion cohort was similar in the blood and liver regardless of which imaging time point, D3-D5, was performed. (**Supplementary Figure 4A-B**)

Figure 3A-B exemplifies the diagnostic tracer’s potential utility in a patient diagnosed with transformed NEPC histology. This patient (#14) was initially diagnosed via a biopsy of the prostate that showed a Gleason 5+5 adenocarcinoma, with focal positive staining of neuroendocrine markers (synaptophysin and chromogranin), and a PSA level of 18. This patient had liver and skeletal metastases at diagnosis and was initially treated with androgen deprivation therapy. However, progressive liver metastases were noted on a follow-up radiological scan only a month later. The patient subsequently had a [^18^F]F-PSMA PET scan that found most lesions to be non-PSMA-avid. A new biopsy of a liver metastases was performed that showed a NEPC histology with DLL3 protein expression (IHC H-score of 175). [^89^Zr]Zr-DFO-SC16.56 PET imaging revealed that most of the patient’s tumor sites (including skeletal, liver, lymph nodes) were strongly avid for the [^89^Zr]Zr-DFO-SC16.56, with the highest uptake found in L3 vertebrae (SUVmax 9·6), and a liver metastasis (SUVmax 8·4). The patient was continued on an ADT, together with carboplatin and etoposide, targeting the NEPC phenotype.

**Figure 3:**
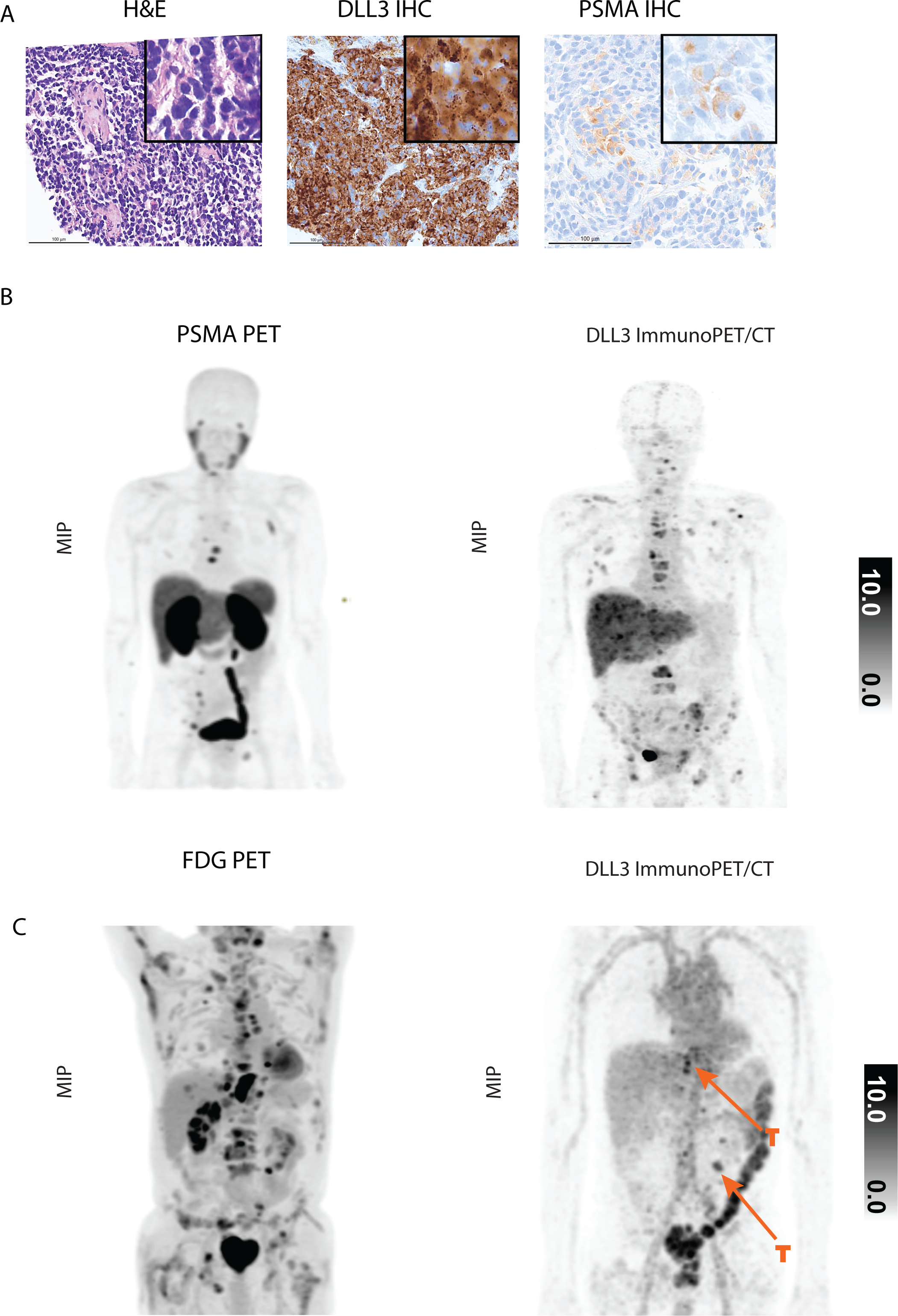
Transformed NEPC (#18) initially diagnosed with prostate adenocarcinoma. The patient had a rapid progression of liver metastases, that were PSMA-non avid **(B)**, with corresponding low PSMA protein expression observed on the IHC of the re-biopsied liver metastasis **(A)**. Furthermore, the liver biopsy showed high-grade neuroendocrine carcinoma, with high DLL3 protein expression **(A).** The DLL3 PET showed that most metastases were DLL3-avid **(B)**. In **Figure 4C**, another NEPC case (#9) demonstrated that of numerous FDG-avid metastases (FDG PET/CT, left), only five metastases were DLL3-avid (marked with T). SUV display scale, pixels in PET imagery with SUV 0.0 appear white and pixels with SUV ≥ 15.0 appear black.

Another NEPC case (#9) showed significant tumoral heterogeneity between the [^18^F]FDG PET/CT and the DLL3 immunoPET/CT (Figure 3C). The patient had previously been treated with androgen deprivation, AR inhibitor, single chemotherapeutic agents (docetaxel and cabazitaxel), as well as four cycles of [^177^Lu]Lu-PSMA. Upon the progression on antiandrogen therapy, a new biopsy of the left perinephric metastatic lesion was performed that showed a transformed NEPC phenotype. The patient was subsequently switched to platinum monotherapy and had response in the bone and lymph nodes after 3 cycles, while liver metastases were stable. After seven cycles of carboplatin, the patient was imaged by DLL3 immunoPET scan, which demonstrated five metastases as DLL3-avid, including the left perinephric metastatic lesion that was biopsied (SUVmax 9·5). The rest of the metastatic lesions were DLL3 non-avid (liver SUVaverage 3·3).

Another case (#12) highlights a transformed NEPC case that had previously received androgen deprivation, AR inhibitor therapy, and showed radiological progression of a liver metastasis that upon re-biopsy showed NEPC transformation. The patient received carboplatin and etoposide, but had rapid disease progression in the liver and was identified as a candidate for a phase I study with a DLL3-targeting T cell engager. Prior to enrollment, the patient was imaged with [^89^Zr]Zr-DFO-SC16.56, which demonstrated no uptake in any of the metastatic lesions (**Supplementary Figure 5**). Analysis of the liver biopsy confirmed low tumoral DLL3 expression (IHC H-score 35). Given these results, the patient was directed toward an alternative therapy and not treated with the DLL3-targeting agent.

## Discussion

In this study, we present a first-in-human trial assessing [^89^Zr]Zr-DFO-SC16.56 as a novel radioimmunoconjugate for *in vivo* imaging of neuroendocrine cancers. The data presented in this study demonstrate the feasibility of DLL3-directed immunoPET/CT imaging of patients with various neuroendocrine cancers, including SCLC and NEPC. These findings are in accordance with preclinical data demonstrating that [^89^Zr]Zr-DFO-SC16.56 can delineate DLL3-expressing tumors in both SCLC and NEPC tumor models.(14, 24) The activity of ^89^Zr used here (nominally 2 mCi) was safe, and no higher than grade 1 toxicities from the radioconjugated antibody were observed. Image quality was deemed satisfactory, making co-injection of cold antibody unnecessary.

Ultimately [^89^Zr]Zr-DFO-SC16.56 as a novel diagnostic tracer has potential to guide treatment selection and stratify patients according to the real-time biological phenotype of the disease, rather than relying on biopsies, which can be associated with significant morbidity and may not be reflective of the totality of disease due to tumor heterogeneity. The approach of assessing DLL3 target expression in a non-invasive manner appears feasible. As the DLL3-targeting agents in clinic trails are expanding, there will be an increased demand for stratifying enrollment by target expression or focusing therapeutic development on those patients most likely to benefit from these therapies.

Striking intrapatient intertumoral heterogeneity was observed when utilizing the radioimmunoconjugate in transformed NEPC patients. Across these cases, imaging by [^89^Zr]Zr-DLL3 PET appeared inversely correlated with [^18^F]F-PSMA– the former presumably detecting transformed high grade neuroendocrine lesions and the latter residual prostate adenocarcinoma. Considering the challenges of conducting multiple biopsies, a non-invasive strategy for detecting molecular features of transformation to NEPC could have substantial clinical utility. This diagnostic tracer could also have potential to monitor the transformation of the CRPC into NEPC to optimize the timing of therapeutic interventions, and to serve as a surrogate endpoint for investigational targeted therapies aimed at treating or constraining transformation.(25)

The radioimmunoconjugate targeting DLL3 can be readily converted to a radiotherapeutic by substituting the imaging radioisotope, ^89^Zr, with a higher energy therapeutic radionuclide (e.g. Lutetium-177) for direct treatment of DLL3-expressing tumor metastases. This strategy has demonstrated efficacy in several preclinical tumor models for SCLC and NEPC and would be the first radiotheranostic approach utilized for these indications.(13, 26) This radiotheranostic strategy has proven successful for other targets, such as in metastatic castration-resistant prostate cancer targeting the prostate-specific membrane antigen (PSMA). FDA approval for the imaging and therapeutic agents [^68^Ga]Ga-PSMA-11 and [^177^Lu]Lu-PSMA-617 can be used as a benchmark for the potential reach of a radiotheranostic program for the DLL3 target.(27, 28)

Weaknesses of this study include the limited number of patients imaged, and that detailed dosimetry was performed on only 3 patients. However, tumor imaging by [^89^Zr]Zr-DFO-SC16.56 was confirmed in 15 of 16 patients with DLL3 H-score of at least 100 and specificity was supported by absence of tumor imaging in the 2 patients with exceptionally low protein expression. Future studies may also assess dynamic changes in DLL3 immunoPET in patients on DLL3-directed therapies, to assess whether target suppression or loss may be a mechanism of acquired resistance to these agents.

## Conclusion

This first-in-human DLL3 immunoPET/CT imaging study demonstrated that [^89^Zr]Zr-DFO-SC16.56 is safe, feasible, and well-tolerated. The data support the ability of [^89^Zr]Zr-DFO-SC16.56 to successfully delineate DLL3-avid tumors in various high grade neuroendocrine cancers. [^89^Zr]Zr-DFO-SC16.56 imaging shows favorable kinetics for early imaging and is associated with intratumoral DLL3 expression assessed by IHC.

## Data Availability

All data produced in the present study are available upon reasonable request to the authors.

## Acknowledgments

We acknowledge support by NIH R01CA213448 (JTP), R35 CA263816 (CMR), U24 CA213274 (CMR), R35 CA232130 (JSL), and a Prostate Cancer Foundation TACTICAL Award (JSL), Scannell foundation. The Radiochemistry and Molecular Imaging Probes Core Facility is supported by NIH P30 CA08748. Funding source was not involved in the study design, data collection, analysis, and interpretation of data; in the writing of the report; and in the decision to submit the paper for publication. We thank the patients for their participation.

---

**Supplementary Table 1:** Inclusion and exclusion criteria evaluated 14 days prior to enrollment onto the trial.

**Supplementary Table 2:** Radiation doses (mGy/MBq) of [^89^Zr]Zr-DFO-SC16.56 to normal organs estimated for Cohort 1.

**Supplementary Table 3:** Biodistribution of [^89^Zr]Zr-DFO-SC16.56 in non-tumor bearing organs calculated for all patients at the respective time points of imaging (D3-D6 post-injection).

