## Supplemental Figures and Tables for "First-in-human imaging with [^89^Zr]Zr-DFO-SC16.56 anti-DLL3 antibody in patients with high-grade neuroendocrine tumors of the lung and prostate"

### Supplementary Figures

**Supplementary Figure 1**
Representative results obtained during analysis of the [$^{89}$Zr]Zr-DFO-SC16.56 final drug product with high speed liquid chromatography (SEC-HPLC) equipped with mass detector **(top)** and radioactivity detector **(bottom)**.

**Supplementary Figure 2**
The maximum intensity projections of each individual case which includes SCLC, atypical carcinoid, NSCLC with neuroendocrine features, and NEPC. SUV scale – lower threshold SUV 0.0 (white pixels), upper threshold SUV 10.0 (black pixels)

**Supplementary Figure 3**
Comparative distributions of absorbed doses for $^{89}$Zr-Pertuzumab (red) and $^{89}$Zr-SC16.56 (black) in selected organs. Error bars denote the standard deviation.

**Supplementary Figure 4**
A) The uptake of $^{89}$Zr-DFO-SC16.56 (SUVaverages) in the blood for the expansion cohort from D3-D5 post-injection (n=13). Error bars denote the standard deviation.
B) The uptake of $^{89}$Zr-DFO-SC16.56 (SUVaverages) in the liver for the expansion cohort from D3-D5 post-injection (n=12). Error bars denote the standard deviation.

**Supplementary Figure 5**
A NEPC patient (#12) initially diagnosed with prostate adenocarcinoma treated with both ADT and ARI, with subsequent radiological progression of liver metastases. A new biopsy of a liver metastasis showed neuroendocrine differentiation with low DLL3 protein expression **(A).** The DLL3 PET scan demonstrated no tumor uptake of [$^{89}$Zr]Zr-DFO-SC16.56 in any of the known metastatic lesions **(B)**.

**Supplementary Figure 1**

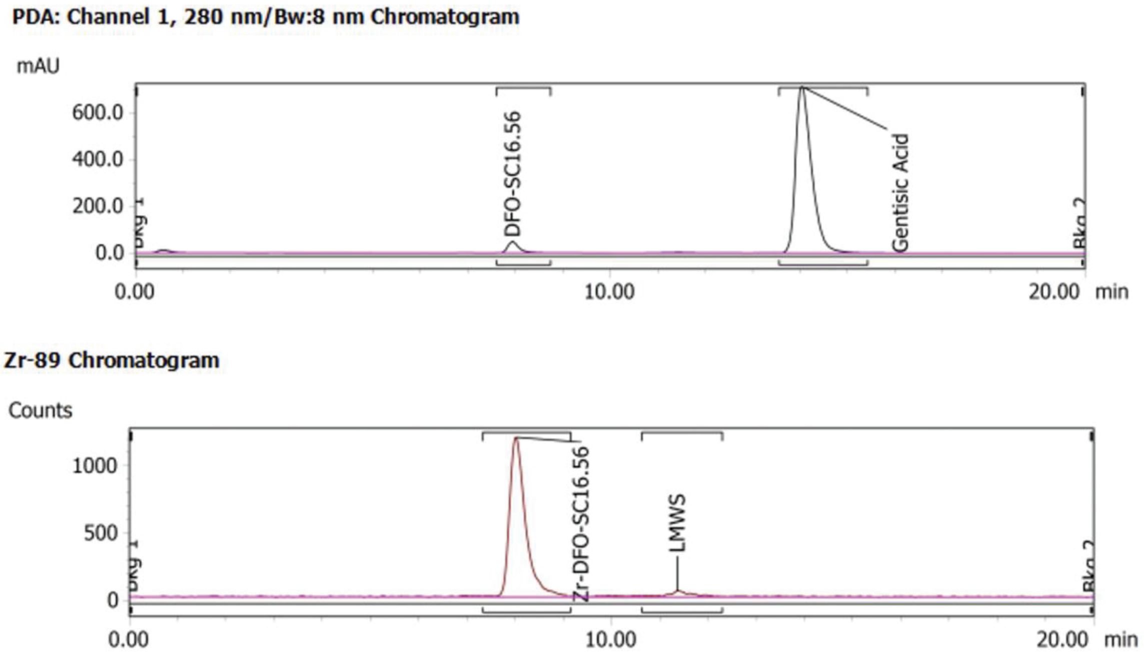

Abbreviations; LMWS: low molecular weight species, mAU: milli-absorbance unit, min: minutes.

**Supplementary Figure 2**

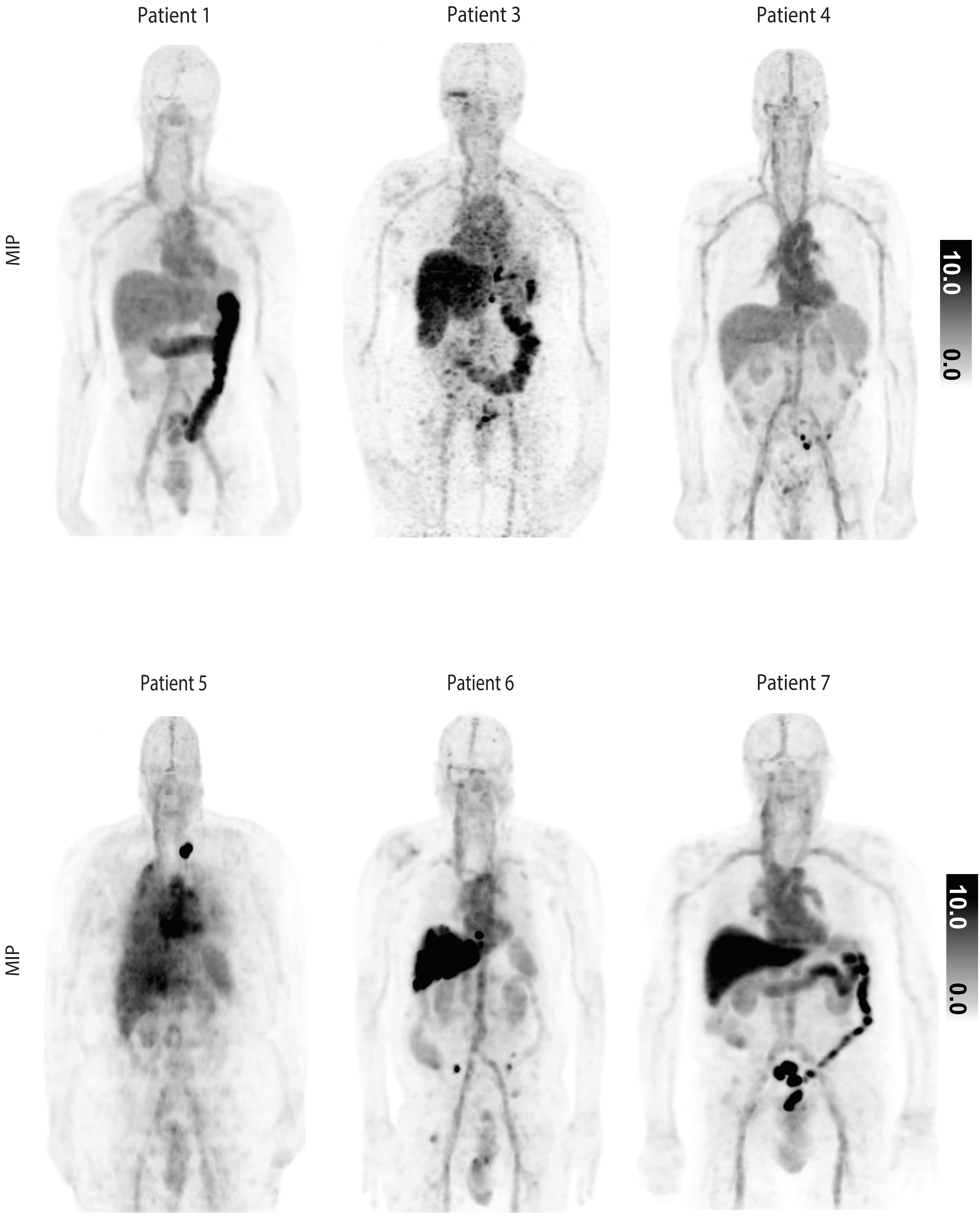

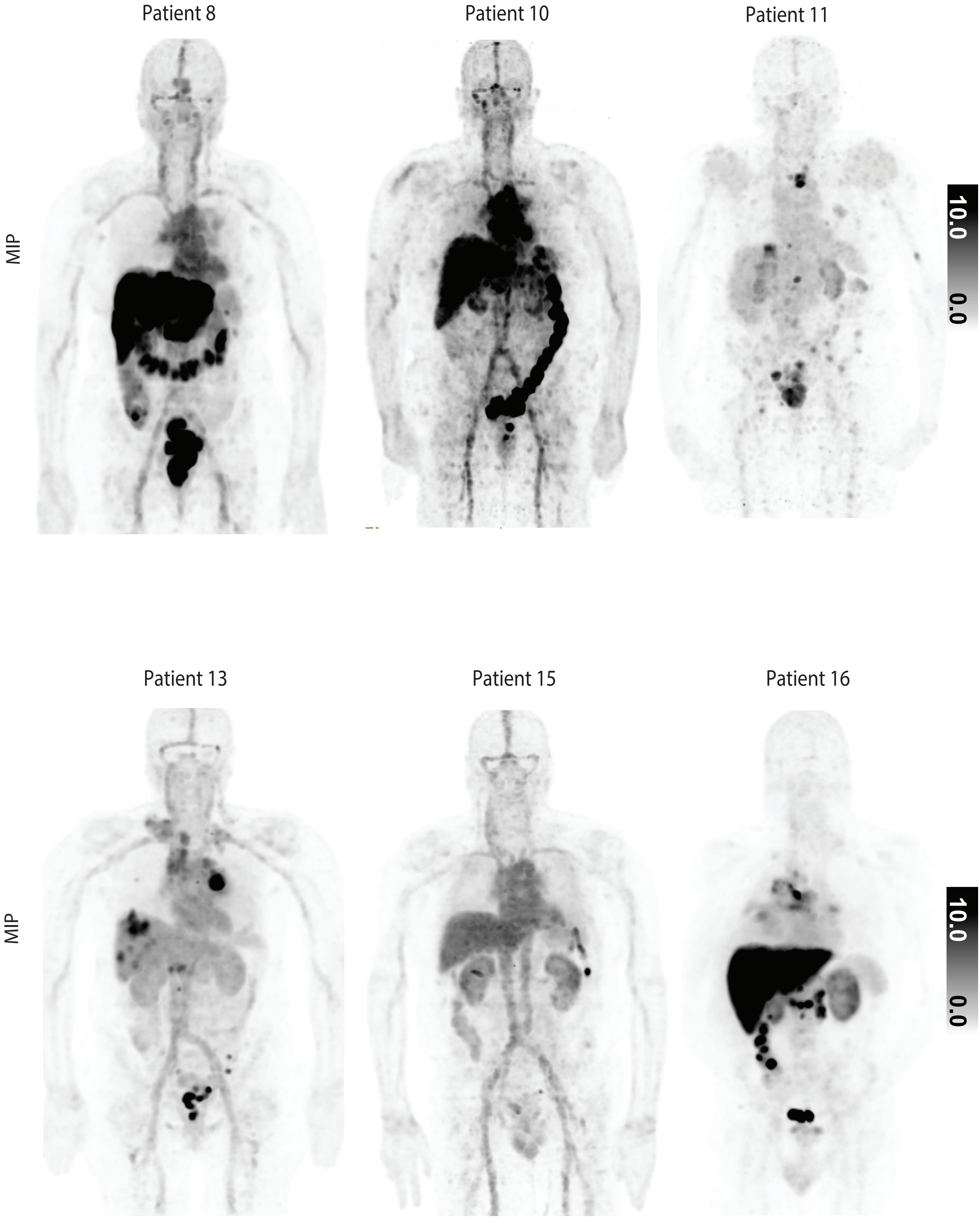
Patient 8
Patient 10
Patient 11
MIP
10.0
0.0
Patient 13
Patient 15
Patient 16
MIP
10.0
0.0

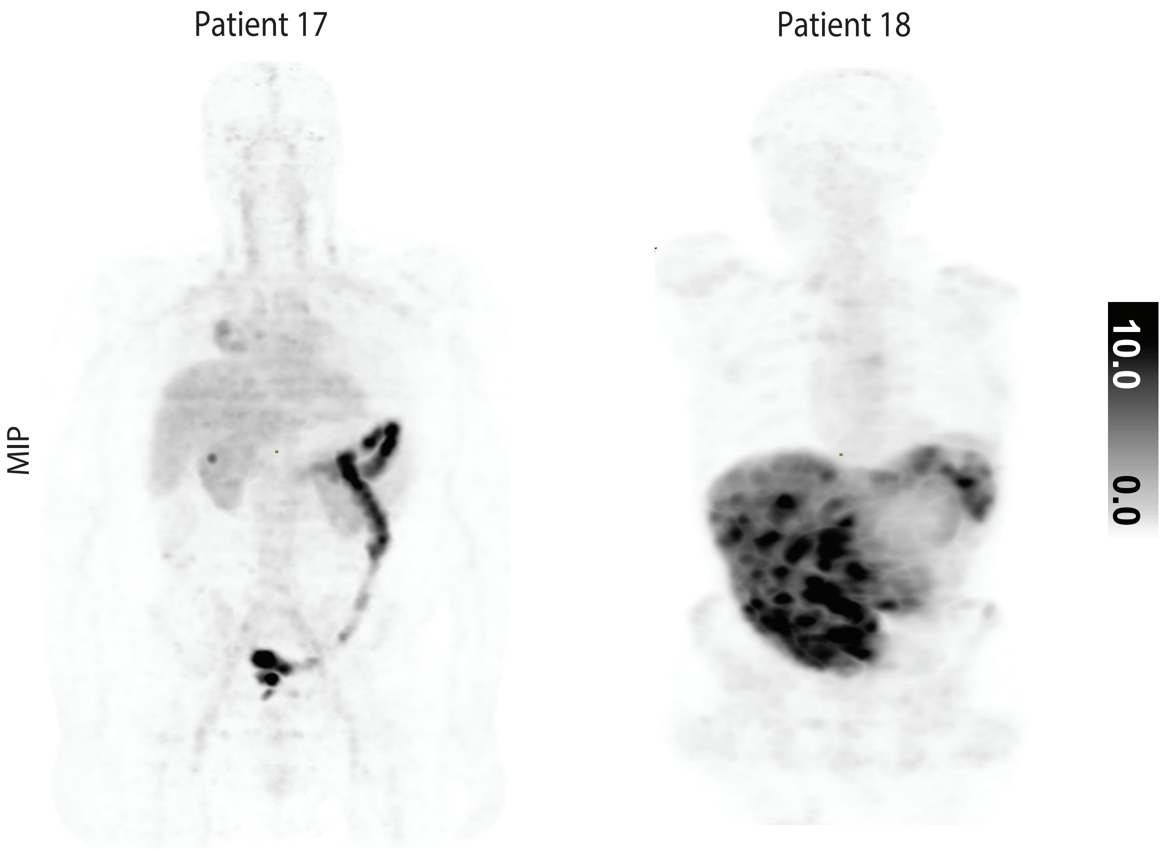
Patient 17
Patient 18
MIP
10.0
0.0

**Supplementary Figure 3**

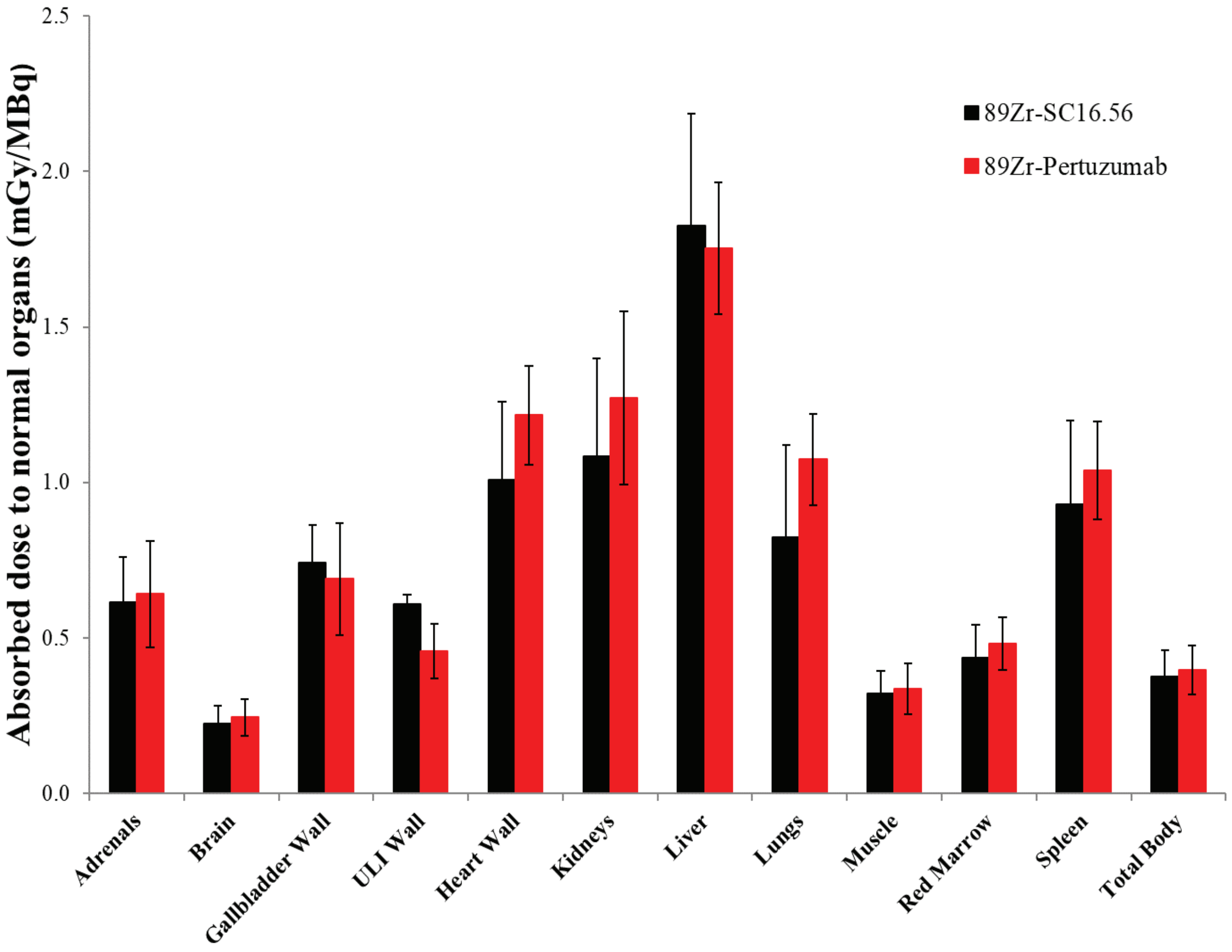

Abbreviations; ULI: upper large intestine

Supplementary Figure 4A

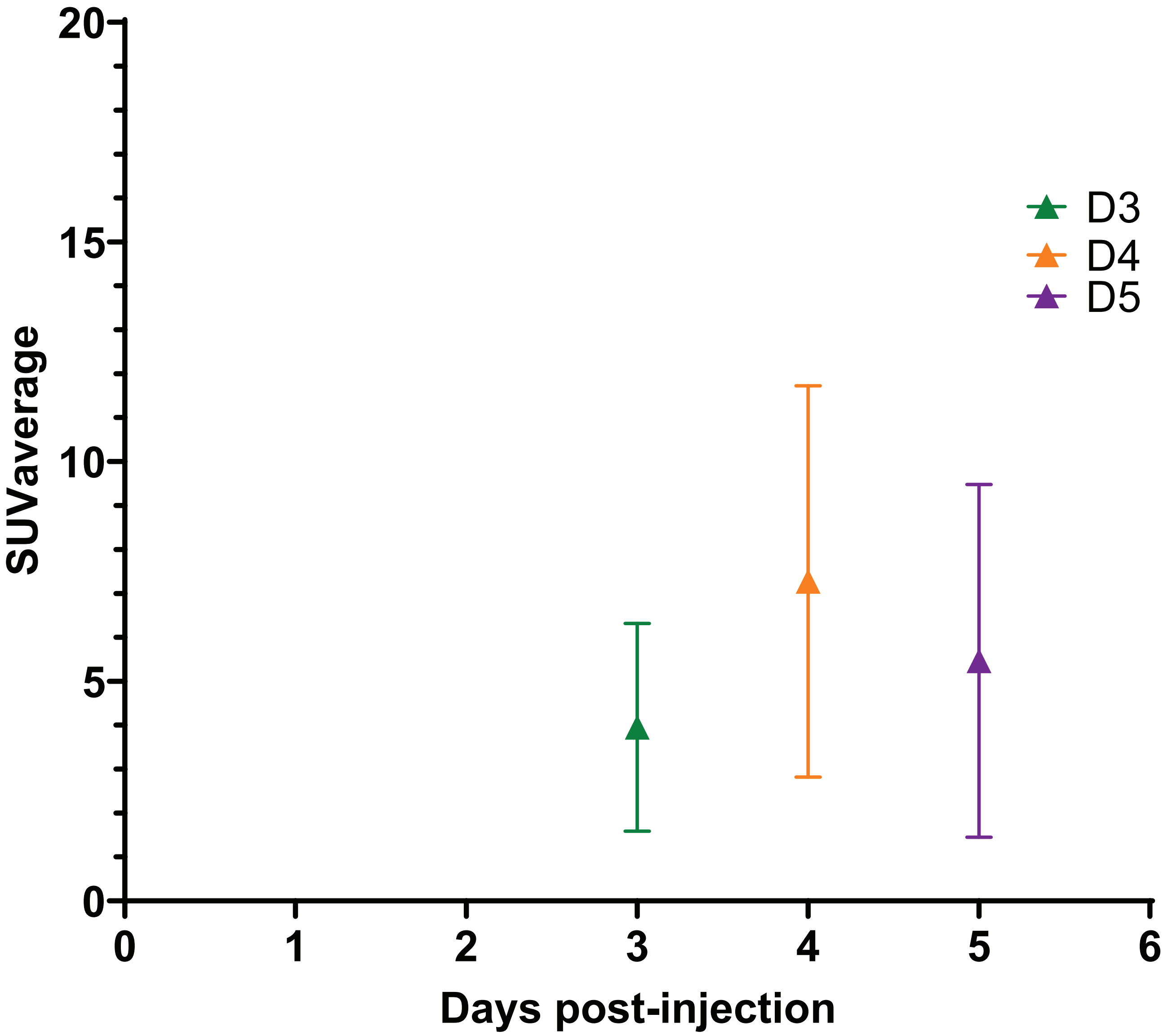

Supplementary Figure 4B

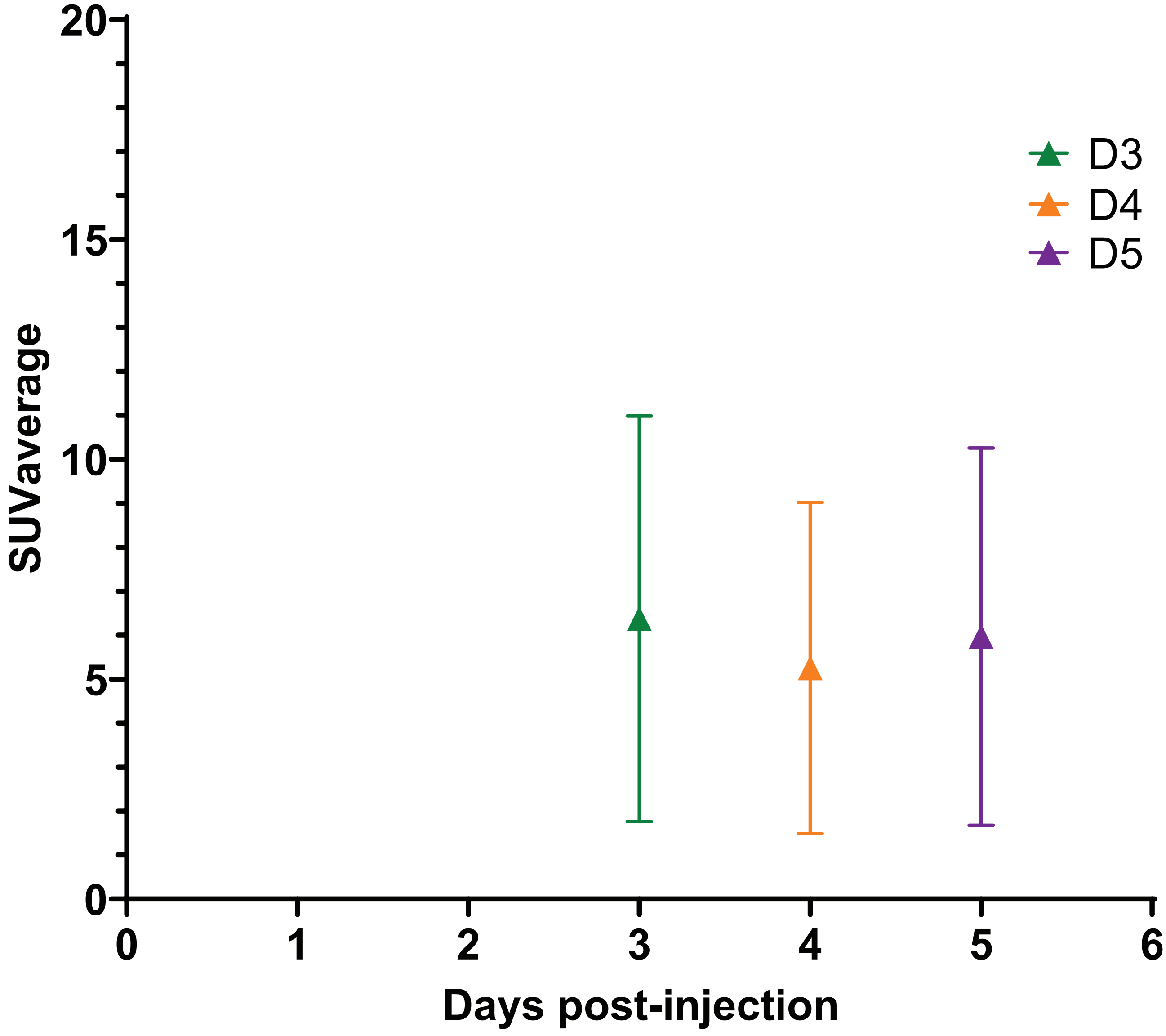

**Supplementary Figure 5**

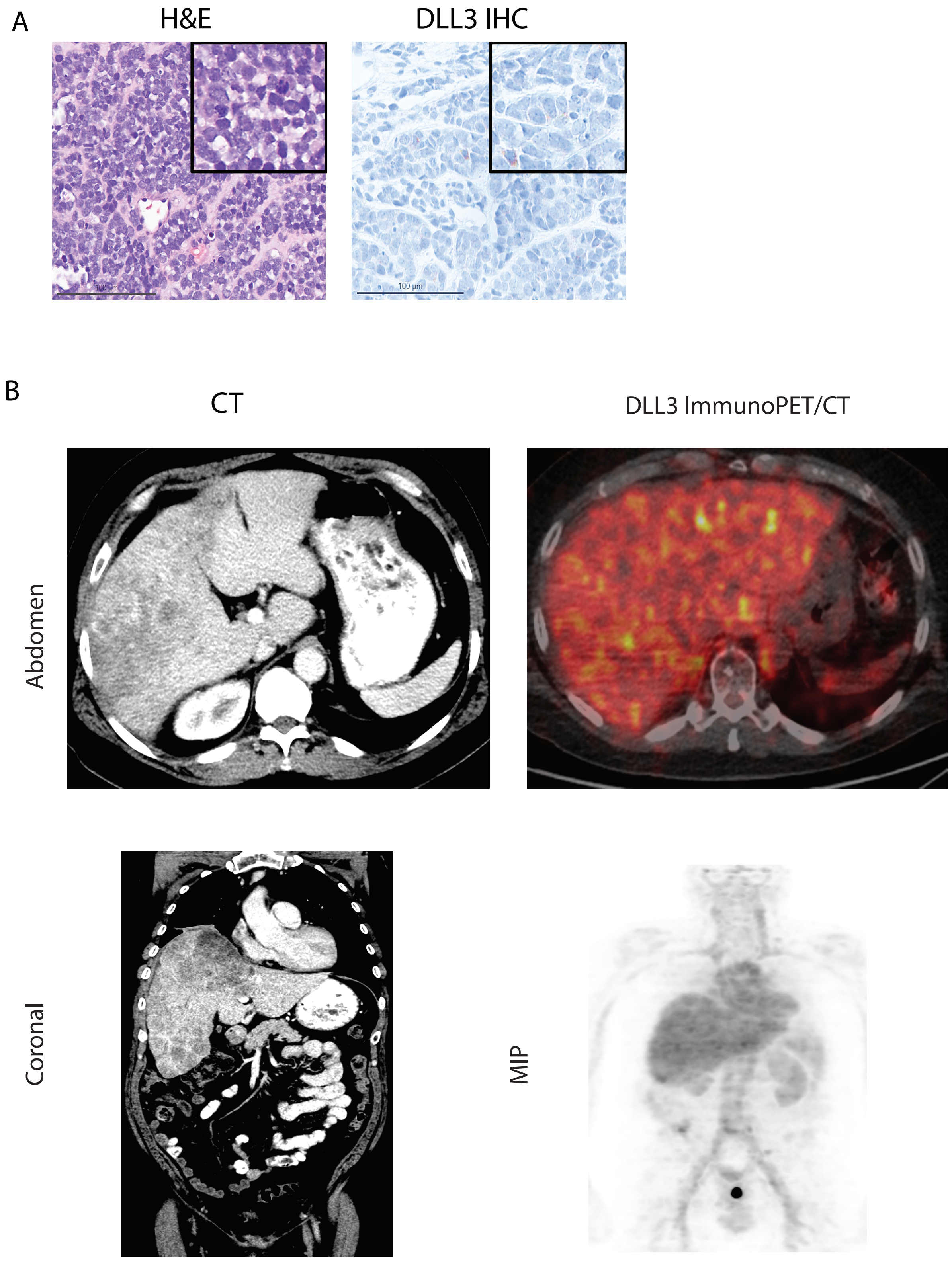

**Table 1**
Demographics and tumor characteristics of Cohort 1 (left) and Expansion cohort (right).

**Table 2**
Patient characteristics for each individual patient included in the study.

**Supplementary Table 1**
Inclusion and exclusion criteria evaluated 14 days prior to enrollment onto the trial.

**Supplementary Table 2**
Radiation doses (mGy/MBq) of [$^{89}$Zr]Zr-DFO-SC16.56 to normal organs estimated for Cohort 1.

**Supplementary Table 3**
Biodistribution of [$^{89}$Zr]Zr-DFO-SC16.56 in non-tumor bearing organs calculated for all patients at the respective time points of imaging (D3-D6 post-injection).

**Supplementary Table 1**

| Subject inclusion criteria | Subject exclusion criteria |
|---|---|
| Patients with histologically verified neuroendocrine derive tumors including SCLC, NEPC, and other histologies. Archival tissue was not required to have been collected within a specific time frame relative to imaging. | History of anaphylactic reaction to humanize or human antibodies. |
| For NEPC patient cohort, rather than archival tumor biopsy material; must have suspected NEPC, based upon clinical assays obtained prior to this trial, and undergo a PET/CT guided biopsy. | Psychiatric illness that would interfere with compliance with the study procedures. |
| At least one tumor lesion measurable on CT or MRI of >/= 1.5 cm. | Pregnant or breast feeding. |
| ECOG Performance status 0 to 2. | Inability to undergo PET scan due to weight limit. |
| Age 18 or more years | |
| Adequate organ function as assessed by:<br><br>Absolute neutrophil count (ANC) ≥ 1·500 $mm^3$<br>Hemoglobin ≥ 8·0 g/dL<br>Platelet count ≥ 75·000/$mm^3$<br>Bilirubin ≤ 1·5 x ULN (upper limit of the norm)<br>AST ≤ 2·5 x ULN (when no liver metastases are present)<br>AST ≤ 5·0 x ULN (when liver metastases are present)<br>ALT ≤ 2·5 x ULN (when no liver metastases are present)<br>ALT ≤ 5·0 x ULN (when no liver metastases are present)<br>Creatinine ≤ 1·5 x ULN | |
| Negative serum pregnancy test within 2 weeks of $[^{89}Zr]$Zr-DFO-SC16.56 for women of child-bearing potential | |

Abbreviations; ALT and AST: alanine aminotransferase and aspartate aminotransferase, ULN; upper limit normal.

**Supplementary Table 2**

| **Patient #** | **1** | **2** | **3** | | | | | |
|---|---|---|---|---|---|---|---|---|
| Sex | M | M | F | | | | | |
| Target Organ | | | | mean | SD | median | min | max |
| Adrenals | 0.59 | 0.49 | 0.77 | 0.62 | 0.14 | 0.59 | 0.49 | 0.77 |
| Brain | 0.21 | 0.17 | 0.29 | 0.22 | 0.06 | 0.21 | 0.17 | 0.29 |
| Breasts | 0.30 | 0.23 | 0.38 | 0.30 | 0.08 | 0.30 | 0.23 | 0.38 |
| Gallbladder Wall | 0.70 | 0.65 | 0.88 | 0.74 | 0.12 | 0.70 | 0.65 | 0.88 |
| Lower Large Intestine Wall | 0.76 | 0.79 | 0.71 | 0.75 | 0.04 | 0.76 | 0.71 | 0.79 |
| Small Intestine | 0.45 | 0.41 | 0.50 | 0.45 | 0.04 | 0.45 | 0.41 | 0.50 |
| Stomach Wall | 0.42 | 0.35 | 0.55 | 0.44 | 0.10 | 0.42 | 0.35 | 0.55 |
| Upper Large Intestine Wall | 0.60 | 0.59 | 0.64 | 0.61 | 0.03 | 0.60 | 0.59 | 0.64 |
| Heart Wall | 1.03 | 0.75 | 1.25 | 1.01 | 0.25 | 1.03 | 0.75 | 1.25 |
| Kidneys | 1.06 | 0.78 | 1.41 | 1.08 | 0.31 | 1.06 | 0.78 | 1.41 |
| Liver | 1.63 | 1.61 | 2.24 | 1.83 | 0.36 | 1.63 | 1.61 | 2.24 |
| Lungs | 0.94 | 0.49 | 1.05 | 0.83 | 0.30 | 0.94 | 0.49 | 1.05 |
| Muscle | 0.31 | 0.26 | 0.40 | 0.32 | 0.07 | 0.31 | 0.26 | 0.40 |
| Ovaries | - | - | 0.48 | 0.48 | - | 0.48 | 0.48 | 0.48 |
| Pancreas | 0.56 | 0.47 | 0.75 | 0.59 | 0.14 | 0.56 | 0.47 | 0.75 |
| Red Marrow | 0.43 | 0.33 | 0.55 | 0.44 | 0.11 | 0.43 | 0.33 | 0.55 |
| Osteogenic Cells | 0.36 | 0.28 | 0.51 | 0.39 | 0.11 | 0.36 | 0.28 | 0.51 |
| Skin | 0.21 | 0.18 | 0.28 | 0.22 | 0.05 | 0.21 | 0.18 | 0.28 |
| Spleen | 0.93 | 0.66 | 1.20 | 0.93 | 0.27 | 0.93 | 0.66 | 1.20 |
| Testes | 0.24 | 0.21 | - | 0.22 | 0.02 | 0.22 | 0.21 | 0.24 |
| Thymus | 0.42 | 0.32 | 0.52 | 0.42 | 0.10 | 0.42 | 0.32 | 0.52 |
| Thyroid | 0.28 | 0.22 | 0.34 | 0.28 | 0.06 | 0.28 | 0.22 | 0.34 |
| Urinary Bladder Wall | 0.32 | 0.28 | 0.33 | 0.31 | 0.03 | 0.32 | 0.28 | 0.33 |
| Uterus | - | - | 0.45 | 0.45 | - | 0.45 | 0.45 | 0.45 |
| Total Body | 0.36 | 0.30 | 0.47 | 0.38 | 0.08 | 0.36 | 0.30 | 0.47 |
| Effective Dose (mSv/MBq) | 0.49 | 0.39 | 0.59 | 0.49 | 0.10 | 0.49 | 0.39 | 0.59 |

Units: mGy/MBq unless otherwise specified.

**Supplementary Table 3**

| Normal organ data (SUV) | SUVaverage | STDEV |
|---|---|---|
| Brain | 0.15 | 0.08 |
| Thyroid | 1.04 | 0.44 |
| Skeletal muscle (Shoulder) | 0.37 | 0.18 |
| Cardiac blood pool | 4.93 | 3.11 |
| Lungs | 0.87 | 0.84 |
| Liver | 5.39 | 3.37 |
| Spleen | 2.85 | 1.37 |
| Pancreas | 1.43 | 1.02 |
| Adrenals | 2.22 | 1.09 |
| Kidneys | 3.16 | 1.05 |
| Large intestine | 1.86 | 1.20 |
| Small intestine | 1.30 | 1.05 |
| Uterus/Prostate | 2.02 | 1.16 |
| Ovaries/Testes | 2.28 | 1.27 |
| Bone (femur) | 0.68 | 0.61 |
| Bone Marrow (Lumbar) | 1.56 | 0.74 |

Abbreviations: SUV: Standardized uptake value, STDEV: Standard deviation.
